# Chronic elevation of serum S100B but not neurofilament-light due to frequent choking/strangulation during sex in young adult women

**DOI:** 10.1101/2021.11.01.21265760

**Authors:** Isabella L. Alexander, Megan E. Huibregtse, Tsung-Chieh Fu, Lillian M. Klemsz, J. Dennis Fortenberry, Debby Herbenick, Keisuke Kawata

## Abstract

Being choked/strangled during a partnered sex is an emerging sexual behavior, particularly prevalent among adolescent and young adult women, but the neurobiological impact of choking remains unknown. This case-control study aimed to test whether frequent choking during sex influences neurological health in young adult women, as assessed by serum levels of S100B and neurofilament-light (NfL). Participants who reported being choking ≥4 times during sex in the past 30 days were enrolled into a choking group, whereas those without were assigned to a control group. Serum samples were collected and assessed for S100B and NfL levels. Demographic questionnaires as well as alcohol use, depression, and anxiety scales were also obtained. Fifty-seven participants were enrolled initially. Due to voluntary withdrawal, phlebotomy difficulties, and scheduling conflicts, the final sample size of 32 subjects (choking n=15; control n=17) was eligible for analysis. After adjusting for a significant covariate (race), the choking group exhibited significantly elevated levels of S100B relative to controls (B=13.96 pg/mL, SE=5.41, p=0.016) but no significant group differences in NfL levels. A follow-up receiver operating characteristic analysis revealed that serum levels of S100B had very good accuracy for distinguishing between the choking and control groups [AUC=0.811, 95%CI (0.651, 0.971), p=0.0033]. Our S100B provide evidence of recurring astrocyte activation due to frequent choking while the NfL data indicate that axonal microstructural integrity may be resilient to these transient hypoxic stressors. Further clinical investigation is needed to clarify the acute and chronic neurological consequences of being choked during sex using a multimodal neurologic assessment.

## INTRODUCTION

Choking a partner during sex has emerged as a prevalent sexual behavior among adolescents and young adults. Our sequential surveys show the prevalence of this potentially harmful sexual behavior, since choking is a form of manual or ligature strangulation that results in obstructed airways and reduced blood flow to the brain.^1^ For example, a recent U.S. nationally representative survey targeting adults ages 18-60 revealed that 21% of sexually active women reported having been previously choked as a part of partnered sexual experiences, as compared to 11% of men.^2^ The prevalence of choking is higher among young adult women and adolescent girls, such that 58% of randomly sampled women college students reported having ever been choked during sex, and one-quarter of these students first experienced being choked between ages 12 and 17.^3^ Further, one in three undergraduate women reported having been choked during their most recent sex.^4^

There are several factors driving the increased prevalence of this sexual behavior, such as influences from pornography, magazines, media, and peers,^2, 5^ which are further complicated by varying perspectives among those who engage in choking during sex. On one hand, choking during sex or erotic asphyxiation is thought to enhance sexual arousal and to be a pleasurable part of sex. In a campus-representative survey in 2020 of 4,989 undergraduate students, a high percentage of respondents (77.2%) considered choking as a form of “rough sex” and 80% of college students who engage in rough sex reported enjoying such sexual behaviors.^1^ On the contrary, some survey respondents have described being choked by a sexual partner as frightening, accompanied by physical and/or emotional distress (e.g., losing consciousness, feeling unsafe).^6^ Moreover, being choked/strangled can be fatal. In addition to deaths associated with partnered sexual asphyxiation,^7, 8^ there have been numerous accidental deaths due to autoerotic asphyxiation,^9, 10^ which is the practice of masturbation while causing oneself to experience cerebral hypoxia to enhance pleasure, often through the use of ligatures or plastic bags.^11, 12^ While epidemiological data on being choked by a partner as a sexual behavior has matured recently, its neurobiological consequences remain completely unknown.

Choking/strangulation can trigger transient ischemic/hypoxic stress and reduced cerebral perfusion that can result in neurovascular uncoupling. In other words, blood flow and its constituents (e.g., oxygen, glucose, metabolites) to the brain becomes insufficient to support neuronal communications and homeostasis. As a result, a glia-mediated inflammatory response emerges. For example, an experimental model of hypoxic-ischemic brain damage in rats demonstrated acute neuronal death and apoptosis of CA1 neurons in the hippocampus following induction of moderate hypoxia.^13^ Furthermore, severe hypoxic-ischemia produced widespread necrosis and infarction.^13^ Similarly, temporary and prolonged blockage of the carotid artery in rats produced permanent damage to the cerebral hemisphere ipsilateral to the occlusion.^14^ While preclinical data about hypoxic-ischemic brain damage is substantive, being choked during sex is a unique human sexual behavior, with varying degrees of intensity, duration, and frequency. Therefore, clinical studies are needed to uncover acute and/or chronic neurologic consequences of being choked during sex.

In pursuit of understanding the neurologic effects of choking, we conducted a case-control study in young adult women undergraduate and graduate students who reported being frequently choked during sexual events (4 times or more in the past month) and a control group of women without reported lifetime experience of being choked. We employed two of the most studied brain-derived blood biomarkers [S100B and neurofilament light (NfL)] to gauge the severity of neurologic distress.^15, 16^ S100B is enriched in astrocytes, and S100B is overexpressed and released by activated astrocytes following cerebral injury and damage.^16^ NfL is a structural protein present in neuronal axons; hence, elevations of NfL levels indicate axonal microstructural damage.^17^ Both S100B and NfL have shown to be elevated in the blood of patients with stroke and traumatic brain injury (TBI), in addition to a subclinical stressor like subconcussive head impacts.^18-21^ However, because choking during sex is a non-mechanical stressors and also a much milder stress than ischemic stroke, we hypothesized that the choking group would exhibit significantly higher serum levels of S100B but not NfL compared to the control group. We further explored the relationship between blood biomarker levels and the frequency of having been choked within the past 30 days, past 60 days, and past 12 months

## METHODS

### Subjects

This case-control study consisted of two groups (choking group vs. control group) and was conducted from February 2021 to June 2021. Potential participants were recruited from our separate campus survey (IRB Protocol # 1912431788A003) and from the university’s online advertisement post. Following consent to study participation, all subjects completed a screening questionnaire to determine eligibility and group assignment. For general inclusion, subjects were required to be birth-assigned female, be enrolled at least part-time at Indiana University, and be between 18 and 30 years old. For the choking group, additional inclusion criteria were that they reported having been choked ≥4 times during partnered sexual events in the past 30 days, whereas women in the control group needed to have been free of any lifetime experience of being choked during a sexual partnered event. Subjects in both groups were excluded if they were pregnant, had a TBI within the past year, reported a history of more than two TBIs, had any magnetic resonance imaging contraindications (e.g., metal inside body near neck, face, or head; metal intrauterine device; severe claustrophobia), or had a neurological condition (e.g., epilepsy, neurodegenerative disease, aneurysm, tumor, spinal cord injury). After confirming eligibility and group assignment, those who qualified for the study were scheduled for data collection (see Fig 1). The Indiana University Institutional Review Board approved the study (IRB Protocol # 10045) and written informed consent was obtained.

**Figure 1.**
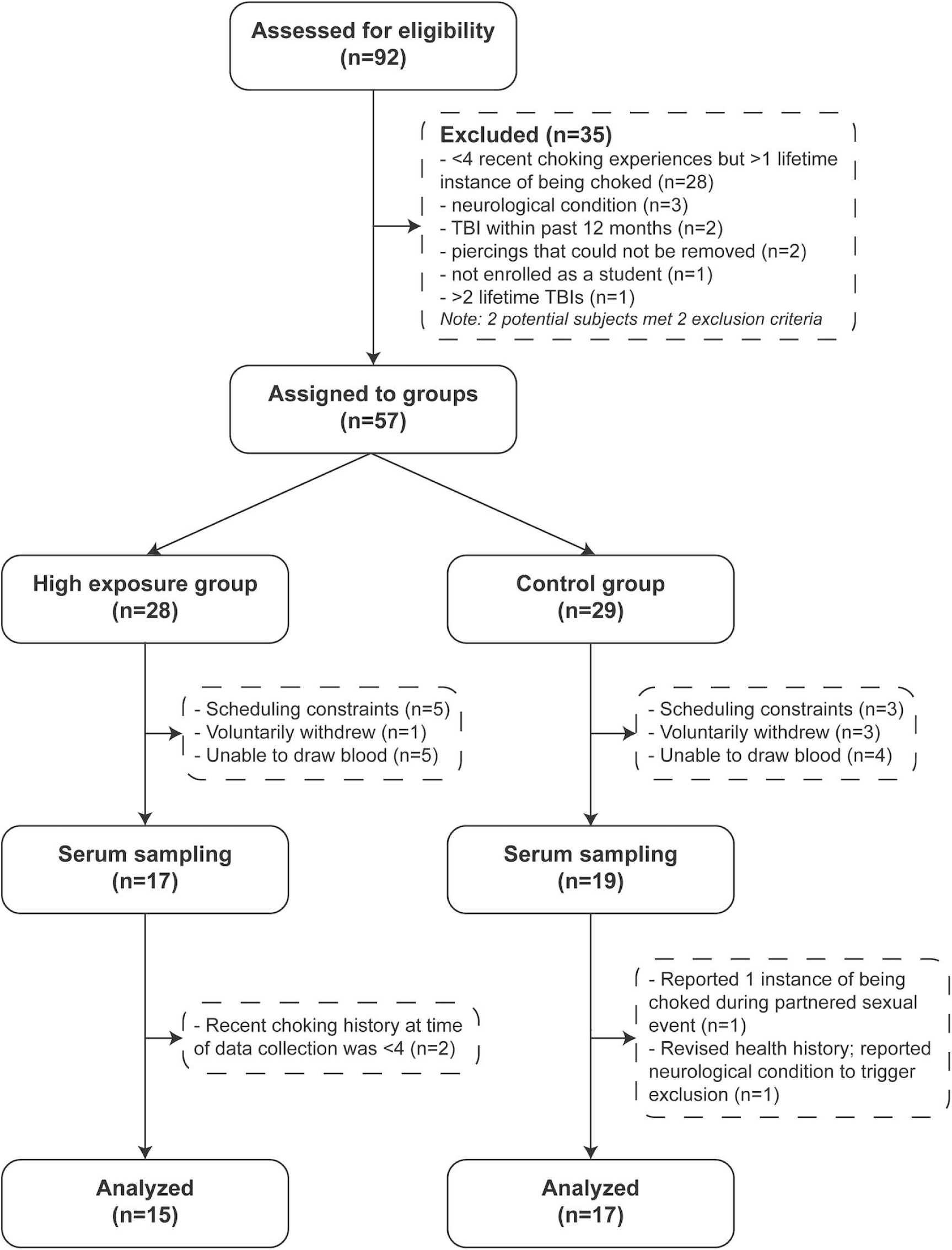
Study Flow Chart.

### Study procedures

Subjects met with one of the two data collection researchers who were blinded to group assignment. Five-milliliter samples of venous blood were collected from the antecubital region into sterile serum vacutainer tubes (BD Biosciences, Franklin Lakes, NJ). Blood samples were allowed to clot at room temperature for a minimum of 30 minutes. Serum was separated by centrifugation (1,500 g, 15 minutes, 4°C) and stored at -80°C until analysis. In addition to completing a questionnaire about their health history and experiences of being choked during partnered sexual events, subjects in both groups completed paper versions of the following mental health scales.

#### Patient health questionnaire (PHQ) for depression and anxiety disorder

Depression-related symptoms were assessed using the depression module of the Patient Health Questionnaire (PHQ-9).^22, 23^ Each of the nine PHQ-9 depression items describes one symptom corresponding to one of the nine Diagnostic and Statistical Manual of Mental Disorders, Fourth Edition diagnostic. Generalized anxiety disorder was assessed using the Generalized Anxiety Disorder Assessment (GAD-7).^24, 25^

#### Alcohol Use Disorders Identification Test (AUDIT)

The AUDIT^26, 27^ is a 10-item screening tool developed by the World Health Organization (WHO) to assess alcohol consumption, drinking behaviors, and alcohol-related problems.

### Biomarker analysis

Serum S100B concentrations were measured using an enzyme-linked immunosorbent assay (ELISA) kit (EMD Millipore Corporation). The lower detection limit of the assay is 2.7 pg/mL using a 50 μL serum sample size, and the assay covers a concentration range of up to 2000 pg/mL, with an inter-assay variation of 1.9–4.4% and an intra-assay variation of 2.9–4.8%. Samples were loaded in duplicate into the ELISA plates according to manufacturer instructions. Fluorescence was measured by a microplate reader (BioTek EL800, Winooski, VT) and converted into pg/mL as per the standard curve concentrations. The S100B ELISA was performed by a member of the research team blinded to the group assignment information.

Serum NfL concentrations were measured using the Simoa^®^ SR-X Biomarker Detection System (Quanterix™, Lexington, MA), a magnetic bead-based, digital ELISA that allows detection of proteins at subfemtomolar concentrations, and an analytical protocol as previously described in detail.^20^ The limit of detection (LOD) for the Simoa^®^ NF-L SR-X assay is 0.0552Lpg/mL, and the lower limit of detection (LLOQ) is 0.316 pg/mL. The Simoa^®^ assay was performed by a certified laboratory personnel blinded to group assignments. Serum samples from all subjects were assayed in duplicate, on the same plate. The average intra-assay coefficient of variation for the samples was 8.2% (standard deviation [SD] 10.3).

### Statistical analysis

The demographic differences between the choking and control groups were assessed by two-sample t-tests and chi-square tests. We used two series of analysis to compare S100B and NfL levels between groups. First, two-sample t-tests were used to assess the group differences in biomarker levels. Second, if t-tests identified a significant group difference, a linear regression was conducted to validate the group difference after adjustment for appropriate covariates. We employed backwards stepwise linear regression using Akaike information criteria (AIC) to determine which covariates to include in the final model. The full (saturated) model included group assignment as the primary predictor with age, race, PHQ-9 scores, GAD-7 scores, and AUDIT scores included as covariates. As a result, race was identified as a significant covariate and therefore included in the final model for the relationship between group assignment and serum S100B. This is in line with previous studies that Black/African American and Asian populations tend to express higher levels of S100B at baseline compared to those who identify as white.^28, 29^

Biomarkers with a significant group difference were further assessed for their diagnostic utility using a receiver operating characteristic (ROC) analysis, and an estimate of the area under the curve (AUC) with 95% confidence interval was obtained. An AUC of 0.5 indicates no discrimination while an AUC of 1.0 indicates a perfect diagnostic utility. To test the potential associations between biomarkers with significant difference between groups and choking frequency, we conducted an exploratory analysis using a series of linear regression models with biomarker level as the outcome variable and reported choking frequency (past 30 days, 60 days, and 12 months) as the predictors. Analyses were conducted using R (version 4.0.2, with the package “olsrr”) and Prism 9 (version 9.0.1). The significance level was set *a priori* at p=0.05, and all tests were two-tailed.

## RESULTS

### Demographic variables

A total of 92 participants were screened for eligibility, and 57 participants who met inclusion criteria and were free of exclusion criteria were enrolled into either the choking group (n=28) or the control group (n=29). Due to voluntary withdrawal, phlebotomy difficulties, and scheduling conflicts, the final sample size of 32 subjects (choking n=15; control n=17) contributed to the biomarker analysis. See Figure 1 for the study flow and reasons for exclusion.

The participants in the choking group had experienced being choked a median of 7, 15, and 30 times in the last 30 days, 60 days, and 12 months, respectively (Table 1). Significant group differences were observed for age, race, and alcohol use. Specifically, the control group was significantly older than the choking group, and the choking group reported significantly higher scores for AUDIT compared to that of the control group. The choking group included more racially diverse participants compared to the control group (Table 1). One S100B measurement in the choking group was considered an outlier as it was more than three standard deviations above the group mean, and this data point was excluded from analysis.

**Table 1.**
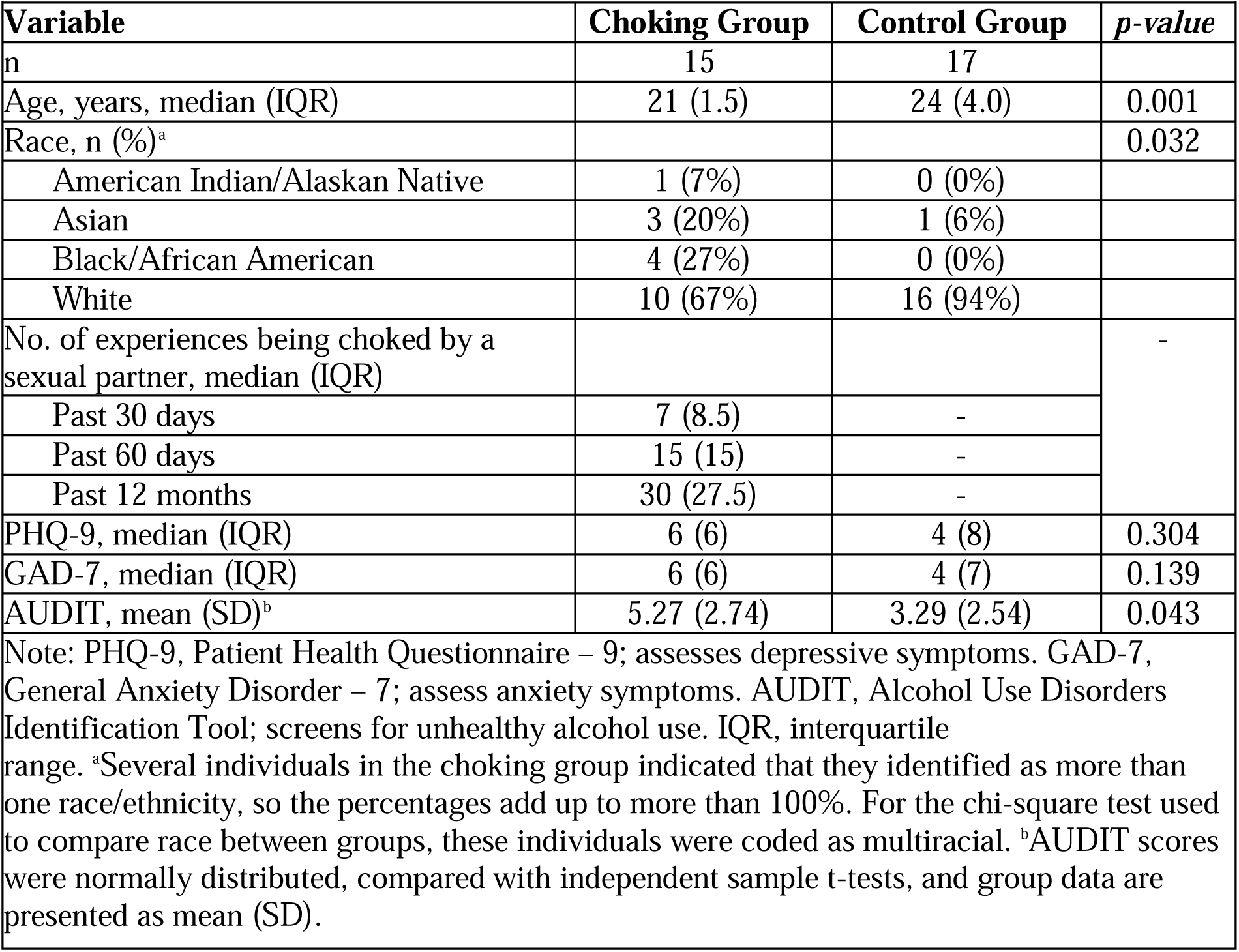
Demographic characteristics.

| **Table 1.** Demographic characteristics. |  |  |  |
| --- | --- | --- | --- |
| **Variable** | **Choking Group** | **Control Group** | ***p-value*** |
| n | 15 | 17 |  |
| Age, years, median (IQR) | 21 (1.5) | 24 (4.0) | 0.001 |
| Race, n (%)<sup>a</sup> |  |  | 0.032 |
| American Indian/Alaskan Native | 1 (7%) | 0 (0%) |  |
| Asian | 3 (20%) | 1 (6%) |  |
| Black/African American | 4 (27%) | 0 (0%) |  |
| White | 10 (67%) | 16 (94%) |  |
| No. of experiences being choked by a sexual partner, median (IQR) |  |  | - |
| Past 30 days | 7 (8.5) | - |  |
| Past 60 days | 15 (15) | - |  |
| Past 12 months | 30 (27.5) | - |  |
| PHQ-9, median (IQR) | 6 (6) | 4 (8) | 0.304 |
| GAD-7, median (IQR) | 6 (6) | 4 (7) | 0.139 |
| AUDIT, mean (SD)<sup>b</sup> | 5.27 (2.74) | 3.29 (2.54) | 0.043 |
| Note: PHQ-9, Patient Health Questionnaire – 9; assesses depressive symptoms. GAD-7, General Anxiety Disorder – 7; assess anxiety symptoms. AUDIT, Alcohol Use Disorders Identification Tool; screens for unhealthy alcohol use. IQR, interquartile range. <sup>a</sup>Several individuals in the choking group indicated that they identified as more than one race/ethnicity, so the percentages add up to more than 100%. For the chi-square test used to compare race between groups, these individuals were coded as multiracial. <sup>b</sup>AUDIT scores were normally distributed, compared with independent sample t-tests, and group data are presented as mean (SD). |  |  |  |

### Group differences and diagnostic utility of blood biomarkers

An initial analysis with two-sample t-tests identified significantly elevated serum S100B levels in the choking group (mean [SD]: 46.00 [15.46] pg/mL) relative to the control group (29.57 [11.11]; p = 0.002; Fig 2A). Conversely, there was no between-group difference in serum NfL levels (Fig 2B). The choking group had elevated serum S100B levels after adjusting for race relative to the control group (B = 13.96, SE = 5.41, p=0.016). A ROC analysis revealed that serum levels of S100B had very good accuracy for distinguishing between the choking and control groups [AUC=0.811, 95% CI (0.651, 0.971), p=0.0033; Fig 3].

**Figure 2.**
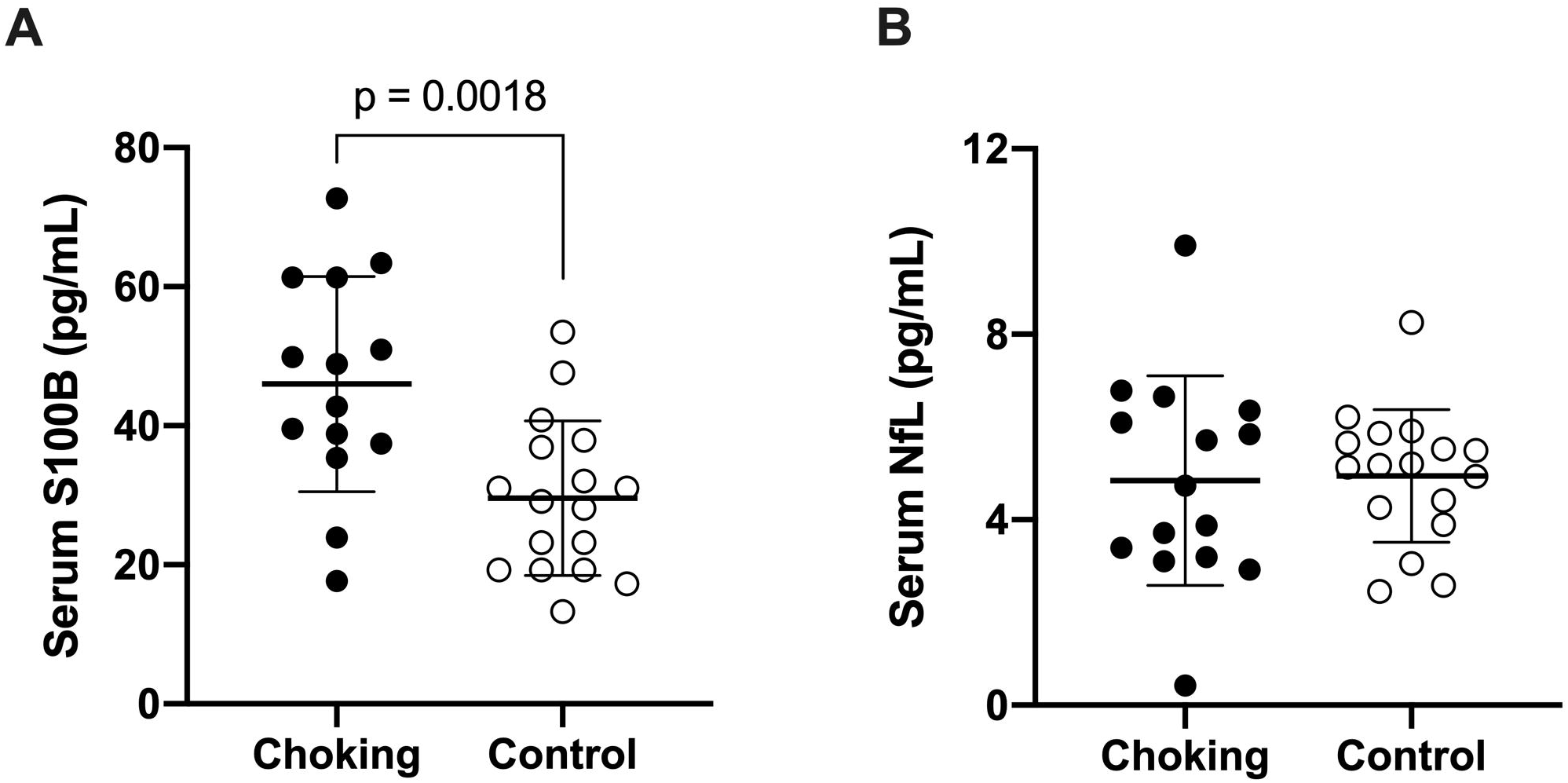
Group differences in serum S100B and NfL levels. The choking group had a significant higher level of S100B compared to that of the control group, whereas NfL levels were comparable between groups. Mean ± SD are depicted by bold lines and error bars. Individual measurements are represented by the closed and open circles.

**Figure 3.**
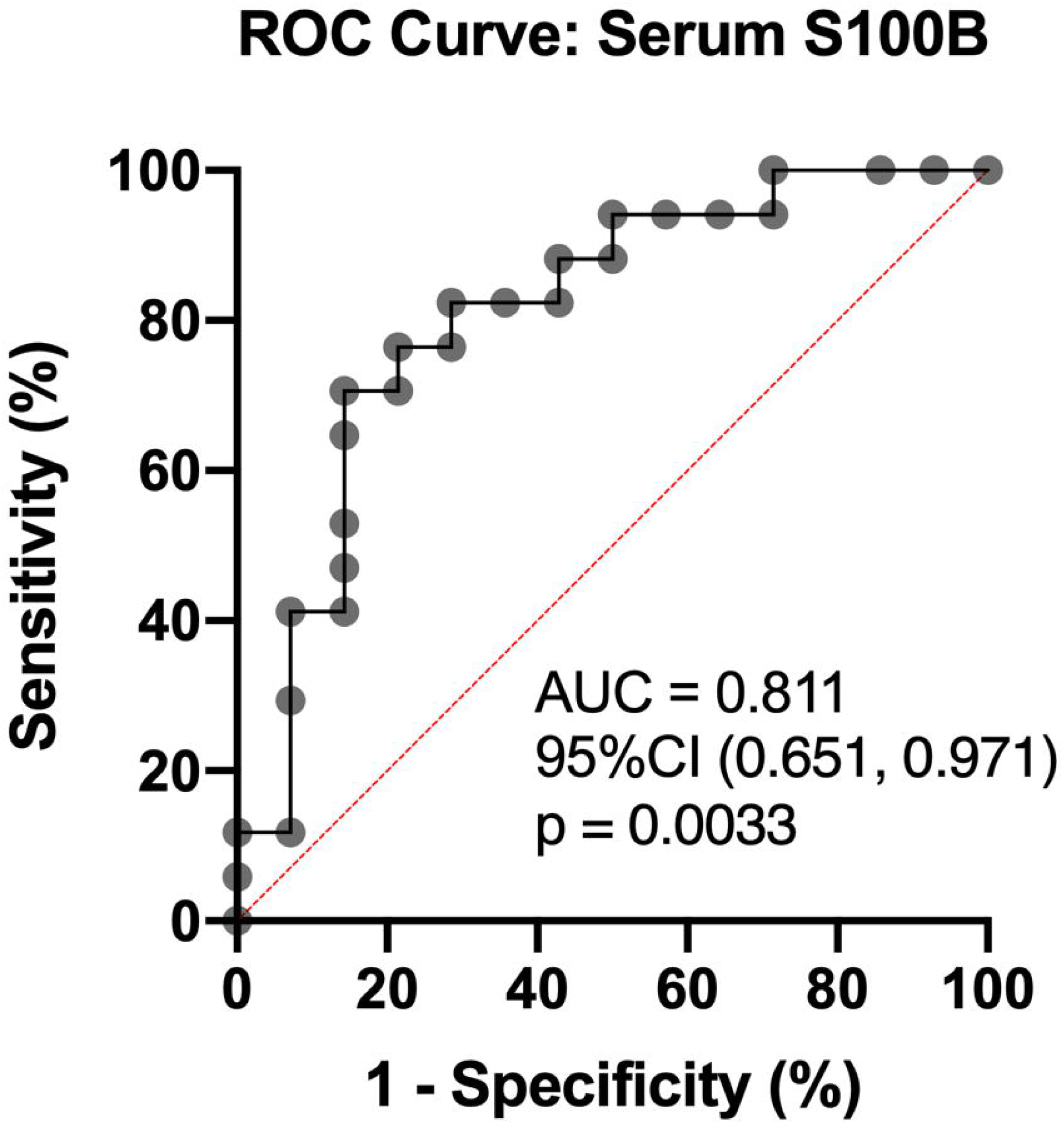
The ROC analysis on S100B. A follow-up ROC analysis revealed that serum S100B had a very good diagnostic accuracy to distinguish the choking group from the control group with an AUC of 0.811, 95% CI (0.651, 0.971), p=0.0033.

### Association between choking frequency and serum S100B levels

A post-hoc analysis was conducted to examine the potential relationship between reported frequency of choking and S100B levels within the choking group. The frequency of choking within the past 30 days, 60 days, or 12 months was not associated with serum S100B levels (Table 2).

**Table 2.**
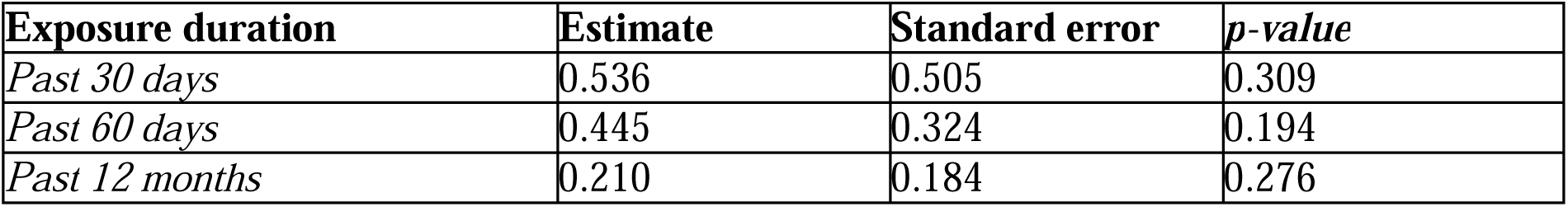
Linear regression model results for the relationships between choking frequency and serum S100B levels.

## DISCUSSION

Within the field of sexual health and behavior research, choking/strangulation as a partnered sexual behavior is a growing area of study with potential clinical and public health implications. While epidemiology studies have revealed the significant prevalence of choking during sex across multiple countries,^3, 30-33^ to our knowledge, this is the first study to investigate the neurobiological consequences of frequent choking during sex using state-of-the-art blood biomarker assays. Our primary finding was that serum S100B levels in women who reported being choked during sex at least 4 times in the past month were significantly elevated relative to choking-naïve controls. Conversely, there was no significant group difference in serum NfL levels. Although choking during sex has been reported to add a euphoric sensation and excitement by young adult and adolescent women,^34^ our data suggest a potential link between frequent choking/strangulation and neuroinflammation that has the potential to ignite meaningful discussion among researchers in sexual health and behavior, neurology, and public health policy.

The novel aspects of the present study include the vulnerable population of young adult women in a case-control design, as well as the use of blood biomarkers shown to be sensitive to repetitive insults to the brain.^18-21, 35^ Our hypothesis was confirmed, as the choking group showed elevated levels of serum S100B but not NfL. Understanding the cellular origin of S100B and NfL and mechanisms required for their overexpression is imperative to interpret our data and contextualize our findings. S100B is enriched in astrocytes, and its primary function is to prevent calcium overload during astrocyte activation due to insults to the brain.^36^ However, the effects of S100B in the brain are concentration dependent. It is protective and neurotrophic at low concentrations but toxic and pro-inflammatory when produced and released by activated astrocytes in high concentrations by activating the receptors for advanced glycation end-product (RAGE) on neuronal cell membranes, triggering NF-κB signal transduction to produce pro-inflammatory cytokines.^36, 37^ The elevated S100B levels in the choking group suggest that frequent choking may provoke astrocyte activation, as well as the astrocyte-mediated inflammatory pathway.

However, it is important to recognize that although S100B showed group differences and elevation in the choking group, S100B elevation was not dependent on choking frequency. The lack of such correlations may be because (1) the relationship between S100B and choking effects may not be linear, (2) there are unknown mediating/moderating factors to the relationship, and/or (3) the recent instances of choking might have influenced the S100B level. In order to address these questions, a follow-up study with a much larger sample size is required. Nonetheless, the existing literature suggests that the degree of S100B elevation is associated with the severity of neurologic stress. For example, S100B levels of the choking group (0.046 ng/mL) are comparable to the levels after sustaining repetitive head impacts from an American football game (0.05 ng/mL).^21^ The serum levels of S100B further elevate in patients with concussion (CT negative, mean 0.18 ng/mL; CT positive, mean 0.36 ng/mL),^38^ severe TBI (with favorable outcomes, 0.3 to 1.6 ng/mL; unfavorable outcomes, 1.1 to 4.9 ng/mL),^39, 40^ and acute stroke (1.1 to 1.4 ng/mL).^41^ This collective evidence suggests that astrocyte activation as reflected by serum S100B levels is sensitive to both mechanical and ischemic-hypoxic stresses to the brain.

Another key finding is that there was no significant group difference in serum NfL levels. NfL is predominantly expressed in long, myelinated axons and has been widely examined for its diagnostic and prognostic utility across TBI, stroke, and neurodegenerative conditions (e.g., dementia, Alzheimer disease).^19, 42^ Unlike traumatic mechanical stress that results in instant axonal disruption, hypoxic stress and cerebral reperfusion effects from choking during sex is transient. Our NfL data indicate that axonal microstructural integrity is resilient to these transient hypoxic stressors, but S100B data provide evidence of recurring astrocyte activation due to frequent choking.

### Clinical and public health relevance

As the topic of choking during sex begins to gain public awareness, it is critical to determine the consequences in terms of brain structure and function of being choked frequently and therefore provide clinicians and other health professionals with sufficient data to generate evidence-based recommendations. It is additionally important to recognize that being choked by a sexual partner may constitute a form of intimate partner violence (IPV) in certain contexts. Globally, 30% of women experience physical and/or sexual IPV in their lifetime,^43^ implicating IPV as a major public health issue with serious ramifications for women’s physical and mental well-being.^43^ While our study is the first to examine levels of brain biomarkers in women who report being choked during partnered sex, several studies have examined chronic effects of IPV-related TBI, which in some cases co-occurred with strangulation.^44-46^ Velara and Kucyi provide evidence that women who sustained TBI as a part of IPV exhibit cognitive deficits that were associated with abnormal functional connectivity and structural integrity.^44^ Additionally, a recent case-series found that IPV victims who reported loss of consciousness showed significant hyperconnectivity in the default-mode network and difficulties with daily functioning and remembering things.^47^ Based on the IPV literature, the next step for the current study is to incorporate a multimodal neuroimaging analysis to examine the choking effects in neuronal function, activation patterns, and microstructural integrity.

### Limitations

There are several key limitations in this study. Our examination of choking was limited to a small sample at a single site tested in a cross-sectional design, limiting the generalizability of the results. A longitudinal study is needed to examine when and how neurologic consequences from choking/strangulation during sex occur and the potential threshold of neurologic resiliency. Self-reported choking behaviors vary in frequency, intensity, and duration, which are subject to recall bias in their survey responses. Moreover, choking during sex is a unique behavior that is not a diagnosable condition, and its operation definition remains subject to interpretation. There is a possibility that elevation in S100B could have occurred due to factors outside of the study protocol. However, because of the choking frequencies reported by subjects, we believe that elevated S100B levels in the choking group were driven by the investigated behavior of choking during sex.

## CONCLUSION

Our data revealed that women who have been choked 4 or more times by a partner during sex in the past month exhibited elevated serum levels of S100B compared to choking-naïve controls, suggesting that transient hypoxia might have provoked astrocyte activation and triggered chronic neuroinflammation. Conversely, axonal microstructural integrity remains resilient to frequent choking during sex, as there was no group difference in serum NfL levels. While this study provides the first empirical evidence of neurobiological consequences of choking during sex, further clinical investigation is needed to clarify the acute and chronic neurological consequences of being choked during sex using multimodal neurologic assessments.

## Data Availability

All data produced in the present work are contained in the manuscript

## Acknowledgements

The authors would like to thank the participants who contributed their time and effort.

## Authorship confirmation statement

KK, DH, JDF, MEH conceptualized and designed the study; ILA, MEH, LMK collected the data; MEH, KK, ILA analyzed the data; ILA, MEH, LMK, TCF, JDF, DH, KK interpreted the data; ILA, KK, MEH drafted the manuscript; MEH, KK, LMK, TCF, JDF, DH critically revised the manuscript for important intellectual content; all authors contributed to the final manuscript and interpretation of the final results.

## Declaration of competing interest

All authors (Alexander, Huibregtse, Fu, Klemsz, Fortenberry, Herbenick, Kawata) report no disclosures relevant to the manuscript.

## Authors’ disclosure statements

I.L.A.: No competing financial interests exist.

M.E.H: No competing financial interests exist.

T.C.F.: No competing financial interests exist.

L.M.K.: No competing financial interests exist.

J.D.F.: No competing financial interests exist.

D.H.: No competing financial interests exist.

K.K.: No competing financial interests exist.

## Funding statement

This publication was made possible with support from the Indiana University School of Public Health (D. Herbenick), the Indiana Clinical and Translational Sciences Institute TL1 Pre-Doctoral Training Award (M.E. Huibregtse; Grant # UL1TR002529 [S. Moe and S. Wiehe, co-PIs], 5/18/2018 – 4/30/2023 from the National Institutes of Health/National Center for Advancing Translational Sciences [NIH/NCATS], Clinical and Translational Sciences Award). This work was also partly supported by National Institutes of Health/National Institute of Neurological Disorders and Stroke (NIH/NINDS)1R01NS113950 (K. Kawata). The funding sources had no role in design or execution of the study; collection, management, analysis, or interpretation of the data; preparation, review, or approval of the manuscript; or decision to submit the manuscript for publication

## Notes

### Competing Interest Statement

The authors have declared no competing interest.

### Funding Statement

This study was funded by Indiana University School of Public Health (D. Herbenick), the Indiana Clinical and Translational Sciences Institute TL1 Pre-Doctoral Training Award (M.E. Huibregtse; Grant # UL1TR002529 [S. Moe and S. Wiehe, co-PIs], 5/18/2018 - 4/30/2023 from the National Institutes of Health/National Center for Advancing Translational Sciences [NIH/NCATS], Clinical and Translational Sciences Award). This work was also partly supported by National Institutes of Health/National Institute of Neurological Disorders and Stroke (NIH/NINDS)1R01NS113950 (K. Kawata). The funding sources had no role in design or execution of the study; collection, management, analysis, or interpretation of the data; preparation, review, or approval of the manuscript; or decision to submit the manuscript for publication.

### Author Declarations

IRB of Indiana University gave ethical approval for this work. IRB Protocol # 10045

